# Detection of HIV and *Mycobacterium tuberculosis* among University Students in Port Harcourt, Nigeria

**DOI:** 10.1101/2021.03.06.21253050

**Authors:** IO Okonko, TI Cookey, EM Okoli

## Abstract

Assessment of HIV and *Mycobacterium tuberculosis* infection is crucial to detect HIV and/or *Mycobacterium tuberculosis* coinfection and the strategy for infection management and treatment. This study assessed the proportion of students with HIV, *Mycobacterium tuberculosis*, and HIV/*Mycobacterium tuberculosis* coinfection. Two hundred and thirty-five university students in Port Harcourt, Nigeria were recruited, ages 16 – 39 years. Samples of blood were collected and processed using standard laboratory procedures. All the students were screened for antibodies to HIV using 2 rapid screening strips and a commercially available enzyme-linked immunosorbent assay (ELISA)-based kit for determination of HIV-1/2/P24/O. The presence of *Mycobacterium tuberculosis* was done using TB rapid kits and a commercially available ELISA-based kit. The results showed that 3.4% of the students were positive for HIV, 2.1% for *Mycobacterium tuberculosis* and none for HIV/*Mycobacterium tuberculosis* coinfection. The age-specific infection rate showed a higher HIV infection rate in the age group 16-24 years (4.7%) than ≥25 years (2.8%). While higher *Mycobacterium tuberculosis* infection rate occurred in the age group ≥25 years (2.8%) than in <25 years (2.3%). The gender-specific infection rate showed that females had a higher infection rate (HIV, 4.7% and *Mycobacterium tuberculosis*, 2.3%) than males (HIV, 2.6% and *Mycobacterium tuberculosis*, 1.7%). Age and sex were the main correlates (P<0.05) of HIV and *Mycobacterium tuberculosis*. This study further confirmed the presence of HIV and *Mycobacterium tuberculosis* infections among University students. These findings suggest the need for regular screening of University students for HIV and *Mycobacterium tuberculosis*.

## INTRODUCTION

Tuberculosis (TB) is an infectious disease caused by *Mycobacterium tuberculosis* (Nazifah et al, 2020), and Africa is facing the worst TB epidemic since the advent of the antibiotic era. Driven by a generalized human immunodeficiency virus (HIV) epidemic and compounded by weak health care systems, inadequate laboratories, and conditions that promote transmission of infection, this devastating situation has steadily worsened, exacerbated by the emergence of drug-resistant strains of *Mycobacterium tuberculosis* (Chaisson and Martinson, 2008; Okonko et al., 2012a,b,c). Detection and control of infectious diseases is a major problem, especially in developing countries (Nazifah et al, 2020).

Human immunodeficiency virus (HIV) and *Mycobacterium tuberculosis* are the two leading causes of death among people living with HIV, accounting for one in four HIV-related deaths (WHO, 2014; Kebede *et al*., 2017), and continue to be a serious problem in developing countries (Kebede *et al*., 2017). At least one-third of the people living with HIV worldwide are infected with latent *Mycobacterium tuberculosis* (WHO, 2014). Indeed, tuberculosis (TB) is one of the deadliest infectious diseases that became a significant public health problem worldwide (Sypabekova et al., 2017). The TB epidemic is on the rise in most countries, including Nigeria. This problem is further compounded by HIV coinfection since one-third of HIV and AIDS-related deaths result from *Mycobacterium tuberculosis* infection (Vihrova *et al*., 2007). Over the past decade, the incidence of *Mycobacterium tuberculosis* has increased in several countries in Africa. Available data suggest that this increase is mainly a result of the burden of HIV infection in those countries.

The epidemic of *Mycobacterium tuberculosis* has infected about one-third of the world’s population creating an adverse impact socially and economically in developing countries (Malen *et al*., 2006). *Mycobacterium tuberculosis* is believed to enhance the progression of HIV to AIDS (Haskins *et al*., 2009). This enhancement of the virus has been suggested to be a result of the generalized immune activation seen in tuberculosis patients (Paton *et al*., 2005; Okonko et al., 2012a,b). Chaisson *et al*. (2004) reported that HIV infection appears to be a key component in the development of active *Mycobacterium tuberculosis* by rapidly increasing its progression. Although very little substantial evidence is currently available, *in vitro* studies have shown that *M. tuberculosis* induces HIV replication increasing the viral load in patients with *Mycobacterium tuberculosis* (Chaisson *et al*., 2004).

While *Mycobacterium tuberculosis* (Mtb) remains the fourth leading cause of death in Africa replacing malaria and other communicable diseases, the number of deaths has dropped by more than half, falling from over 1 million in 2000 to 435,000 in 2019 in Africa (WHO, 2020). *Mycobacterium tuberculosis* is also no longer in the global top 10, falling from 7th place in 2000 to thirteenth in 2019, with a 30% reduction in global deaths (WHO, 2020). However, *Mycobacterium tuberculosis* remains the most common presenting illness among people living with HIV, including those who are taking antiretroviral treatment (WHO, 2014). In the same vein, HIV/AIDS dropped from the 8th leading cause of death in 2000 to the 19th in 2019, reflecting the success of efforts to prevent infection, test for the virus and treat the disease over the last two decades (WHO, 2020). There is a dearth of literature on the seropositivity of HIV and *Mycobacterium tuberculosis* coinfections and associated risk factors among Nigerian University students to the best of our knowledge.

Lateral flow immunoassay (LFIA) has been introduced as a handheld immunoassay-based point-of-care platform for automated detection of TB (Nazifah et al, 2020). Paper-based immunoassays represent a powerful technique with high relevance in biosensing as it achieved the requirements needed for point-of-care (POC) devices, including rapid detection, a small amount of sample, and low cost so that more people can conveniently receive cost-effective healthcare in resource-limited areas (Li et al., 2019; Nazifah et al, 2020). The paper-based POC immunoassays are generally composed of three major components, i.e., paper as the substrate, antibodies as the detection element, and reporter molecules as the signal-transforming element (Nazifah et al, 2020). Lateral flow immunoassay (LFIA), also known as strip-based biosensing, is one of the existing paper-based platforms that represent the most favourable strategy for on-site and one-shot sensor (disposable) analysis (van Pinxteren et al., 2000; Li et al., 2019; Nazifah et al, 2020). It is worth mentioning that electrochemical approaches are also taking advantage of lateral flow strips (Ruiz-Vega et al., 2017). However, LFIA has some drawbacks; for example, at low concentrations of analyte, this technology may present problems in terms of sensitivity (Zamora-Gálvez et al., 2018; Nazifah et al, 2020).

Concerning handheld detection platforms for the POC, particularly for *Mycobacterium tuberculosis* detection, colourimetric-based LFIA appears to be a good start since this method realized rapid diagnostics in the field conditions as well as simple operation (Nazifah et al, 2020). Besides, early detection is so critical to enabling the timely initiation of antituberculosis treatment, which is the key to reduce mortality as well as to avoid the *Mycobacterium tuberculosis* epidemic (Nazifah et al, 2020). To achieve that, we have focused on detecting the presence of HIV and TB infections among University students in Port Harcourt, Nigeria to generate baseline information.

## MATERIALS AND METHODS

### Study Area

The study was conducted among students of the University of Port Harcourt located in Choba, Rivers State, Nigeria. It is a mixed tertiary institution made up of 13 and 55 departments comprising of undergraduate, postgraduate and doctorate students.

### Study Population

A total of two hundred and thirty-five university students of different ages, sex and socioeconomic status presenting at the O. B. Lulu Briggs Health Centre (The University of Port Harcourt Medical Centre) in Port Harcourt, Nigeria was enrolled in this study. The study was conducted by recruiting consecutive consenting students presenting at the health centre for their medical examinations until a total of 235 participants was attained. Other relevant information of all participants was obtained using a Performa specially designed for this purpose. The study was approved by the management of the O. B. Lulu Briggs Health Centre and ethical approval was obtained from the Research Ethics Committee of the University of Port Harcourt, Nigeria. All work was performed according to the International Guidelines for Human Experimentation in Clinical Research. The diagnosis of HIV and *Mycobacterium tuberculosis* infection was established by a standard enzyme-linked immunosorbent assay (ELISA).

### Detection of HIV Antibodies

All the recruited students were screened for antibodies to HIV using 2 enzyme immunoassay (EIA) rapid screening kits, based on WHO systems-2 for detecting antibodies to HIV-1 & 2, and a commercially available enzyme-linked immunosorbent assay (ELISA) kit. For the detection of the presence of HIV-1 and/or HIV-2 antibodies in the blood samples collected, a World Health Organization (WHO) approved kits called ‘DETERMINE® HIV-1/2 (Abbott laboratories) and HIV-1/2 STAT-PAK® (Chembio Diagnostic Systems, Inc.)’, as well as a commercially available enzyme-linked immunosorbent assay (ELISA) based kit (manufactured by Dia. Pro, Milano, Italy), were used. The test was carried out according to the manufacturer’s specifications. Both positive and negative control sera were run along with the test samples using the same procedure. A seropositive test means observable seroreactivities in HIV with both rapid test strips and a commercially available ELISA based kit. No seroreactivity and seroreactivity to a single detection kit was recorded as seronegative.

### Detection of Mycobacterium tuberculosis

#### Lateral flow immunoassay (LFIA) test

“Tuberculosis lateral flow immunoassay (LFIA) also called immunochromatographic strip test” (manufactured by Global Inc., USA), are a chromatographic immunoassay (CIA) for the direct qualitative detection of tuberculosis antibody in human serum or plasma specimens. LFIA test strips are widely used as an aid in the diagnosis of infection due to *Mycobacterium tuberculosis* because this test is easy to perform. Tuberculosis LFIA test cassette employs a unique combination of a specific antibody binding protein conjugated to colloidal gold particles and antigens, which are bound to the membrane solid phase.

#### Conventional Enzyme-Linked Immunosorbent Assay (ELISA) Test

To confirm the positivity and negativity of the samples in comparison to the LFIA used, a semiquantitative ELISA test was done using commercially available TB ELISA based kits (manufactured by Dia. Pro, Milano, Italy). Briefly, ELISA 96-well plates were coated with 100□μl of rabbit anti-*M. tuberculosis* antibody with a concentration of 1□μg/ml in carbonate buffer. Two hundred microliter (200 ul) of Negative Control were dispensed in triplicate and 200 ul Positive Control was dispensed in a single in proper wells. Two hundred microliter (200 ul) of Sample Diluent (DILSPE) was added to all the sample wells; then 10 ul of the sample was dispensed in each properly identified well. The plate was gently mixed, avoiding overflowing and contaminating adjacent wells, to fully disperse the sample into its diluent. Fifty microliters (50 ul) of Assay Diluent (DILAS) were dispensed into all the controls/calibrator and sample wells. The ELISA plate was covered with parafilm and incubated for 45 mins at 37°^C^. Then, each coated well was washed 3 times by filling the wells with 350 ul of washing buffer containing phosphate buffer solution (PBS) and Tween-20. All the solutions were removed by flicking the plate 2-3 times to remove any unbound protein. After the washing step, the ELISA plate was blocked by adding 100μl Enzyme Conjugate into each well, except the 1st blanking well, and covered with the sealer and incubated for 45□min at 37°^C^ room temperature. The ELISA plate was washed again 3 times by washing buffer and 100μl of Chromogen/Substrate mixture was dispensed into each well, the blank well included. Then the microplate was incubated at room temperature (18-24°C) for 15 minutes. Finally, 100□μl of stop solution (0.5□M H2SO4) was added to stop the enzymatic reactions. The addition of acid will turn the positive control and positive samples from blue to yellow. The reading of absorbance at 450nm filter (reading) and possibly at 630nm (background subtraction), blanking the instrument on A1 was obtained by using an ELISA Microplate Reader.

### Interpretation of Results

The mean OD450nm value of the Negative Control (NC) was calculated by applying the following formula: **CUT-OFF = NC + 0.350**. The Sample / Cut-Off value (or S/Co) for the Controls and the samples was also calculated. The value of 0 arbU/ml was assigned to the Negative Control and the value of 100 arbU/ml to the Positive Control. Then on a linear millimetre paper, a line was drawn between the Negative Control and the Positive Control values. S/Co values of samples were then converted into arbU/ml using the curve, providing a semi-quantification of the IgG content in the sample. In the quantitative method, samples showing an OD 450nm value lower than the Cut-Off value were considered negative for anti-MTB IgG. Samples showing an OD450nm value higher than the Cut-Off value were considered positive for anti-MTB IgG. In the Semi-quantitative Method, a quantification of the IgG content in arbU/ml is possible for those samples that show an OD450nm higher than the Cut-Off (or S/C0 > 1); this provides the possibility for the clinician to follow up the immunological status of the tuberculosis patient.

### Data Analysis

All data generated was presented in Tables and subjected to statistical analysis (the χ2-test, with the level of significance set at *p* < 0.05) using statistical package for social sciences (SPSS) to determine any significant relationship between infection rate, age and gender. We calculated trends in reporting rates by age, sex, and department and expressed in average percentage. The seropositivity of HIV and *Mycobacterium tuberculosis* coinfection was determined from the proportion of positive individuals to the total number of the individual under consideration, and it is expressed in percentage.

## RESULTS

### Socio-Demographic Information

Table 1 summarizes the demographic characteristics of students in Port Harcourt, Rivers State, Nigeria used in this study. The age of the students ranged from 16 years to 39 years. One hundred and twenty-eight students (54.5%) were within the age group 16 – 24years while one hundred and seven students (45.5%) were within the age group 25 years and above. Among the students tested, there were more females than males. The females were 120 (51.1%) while the males were 115(48.9%) as shown in Table 1. The ratio of female to male was kept close to 1:1.

**Table 1:**
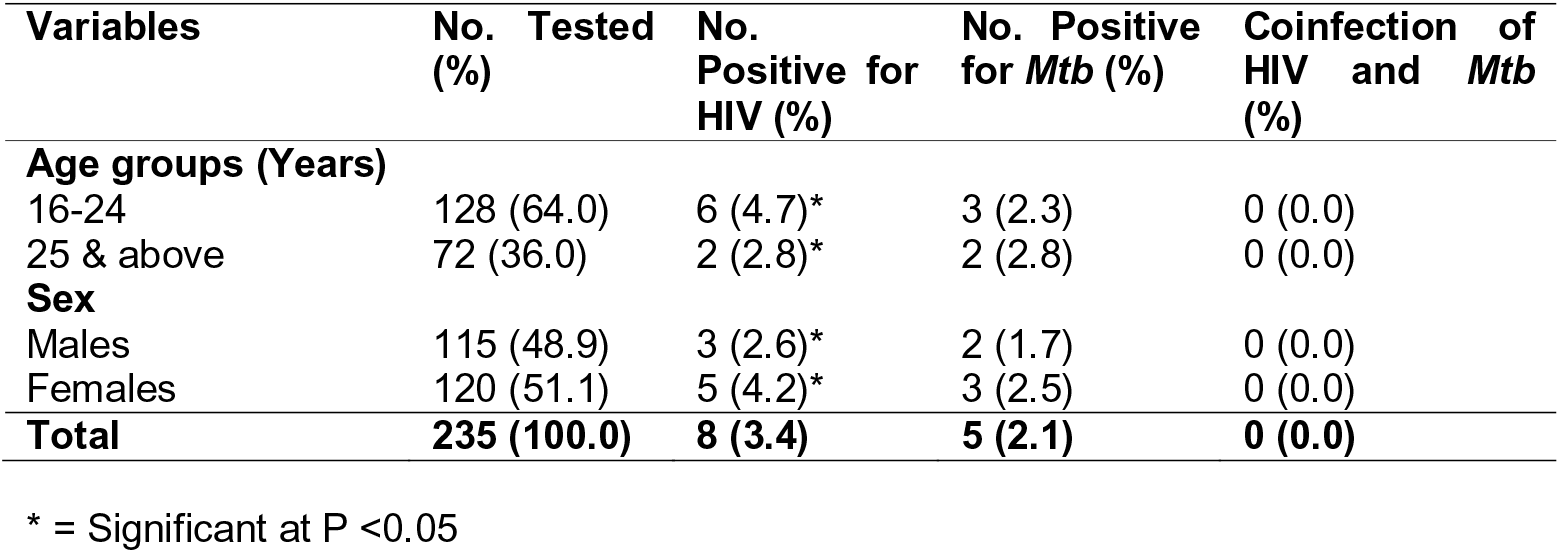
Detection of HIV and *Mycobacterium tuberculosis* (*Mtb*) with socio-demographic characteristics of the students.

### Overall seropositivity of HIV and *Mycobacterium tuberculosis* (*Mtb*)

Of the 235 students who were enrolled on the study, 8(3.4%) were positive for HIV, 5(2.1%) were positive for *Mycobacterium tuberculosis* and none (0.0%) was positive for both *Mycobacterium tuberculosis* and HIV as coinfection. The baseline characteristics of the infected students are shown in Table 1 along with values for the age, sex and socio-economically matched *Mycobacterium tuberculosis* and HIV. Table 1 shows the seropositivity of *Mycobacterium tuberculosis* and HIV with the risk factors.

### Seropositivity of HIV and *Mycobacterium tuberculosis* (*Mtb*) with the ages of students

The age-specific seropositivity showed that students in the age group 16 to 24 years of age had a higher infection rate for HIV (4.7%, n=6) than those in the age group 25 years and above, who had a total of 2(2.8%) infection rate for HIV (Table 1). While students in the age group 25 years and above had a higher infection rate for *Mycobacterium tuberculosis* (2.8%, n=2) than those in less than 25 years (2.3%, n=3). Statistically, it showed a significant difference (P<0.05) in the distribution of infections concerning age.

### Seropositivity of HIV and *Mycobacterium tuberculosis* (*Mtb*) with the sex of students

The sex-specific infection rate showed that females had higher infection rate for HIV (4.7%, n=5) and *Mycobacterium tuberculosis* (2.3%, n=3) than their male counterparts (2.6%, n=3 for HIV and 1.7%, n=2 for *Mycobacterium tuberculosis*). However, there was a significant difference (P<0.05) in the distribution of HIV and *Mycobacterium tuberculosis* concerning sex as shown in Table 1.

### The proportion of students with HIV and *Mycobacterium tuberculosis* (*Mtb*) infection

Out of the two hundred and thirty-five students tested, eight (3.4%) of the students were HIV positive (Figure 1). Of these eight HIV positive students, 5 (62.5%) were females and 3 (37.5%) were males. Of the 235 students tested, 5 students (2.1%) were seropositive to *Mycobacterium tuberculosis* (Figure 2). Of the 5 *Mycobacterium tuberculosis* seropositive students, 2 (40.0%) were males while 3 (60.0%) were females.

**Figure 1.**
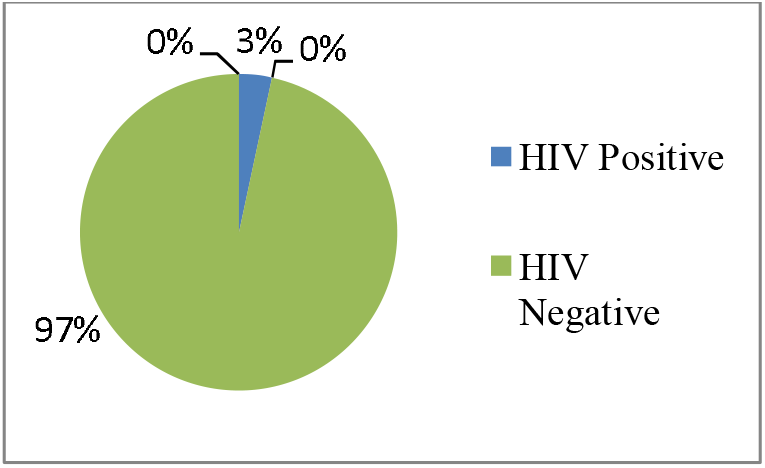
Percentage of Students with and without HIV infection.

**Figure 2.**
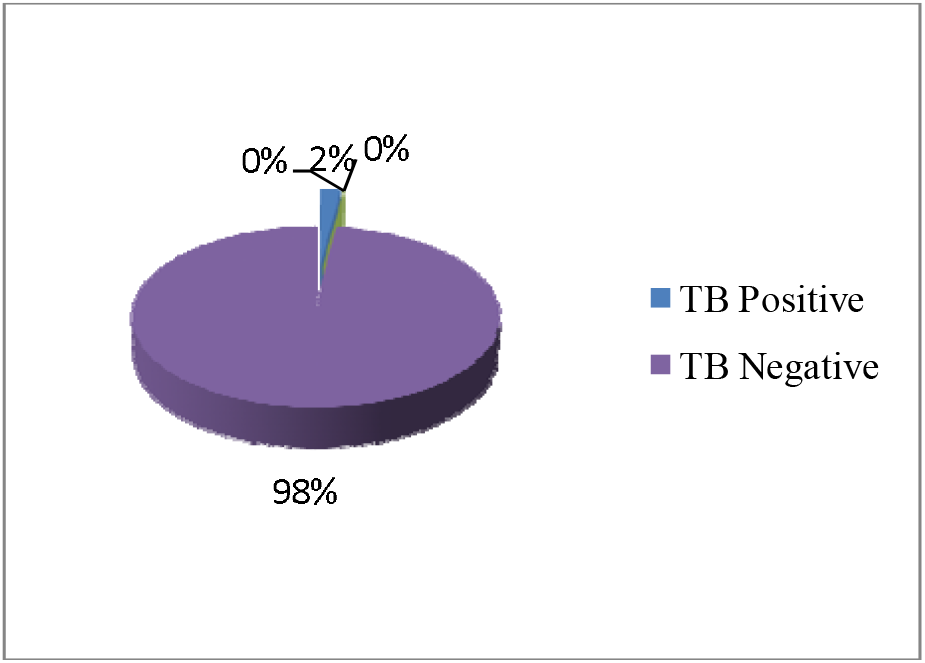
Percentage of Students with and without *Mycobacterium tuberculosis* infection.

### HIV and *Mycobacterium tuberculosis* (*Mtb*) coinfection

Of the eight HIV positive students tested for *Mycobacterium tuberculosis* coinfections, none of the students was co-infected with HIV and *Mycobacterium tuberculosis* (Table 1, Figure 3).

**Figure 3.**
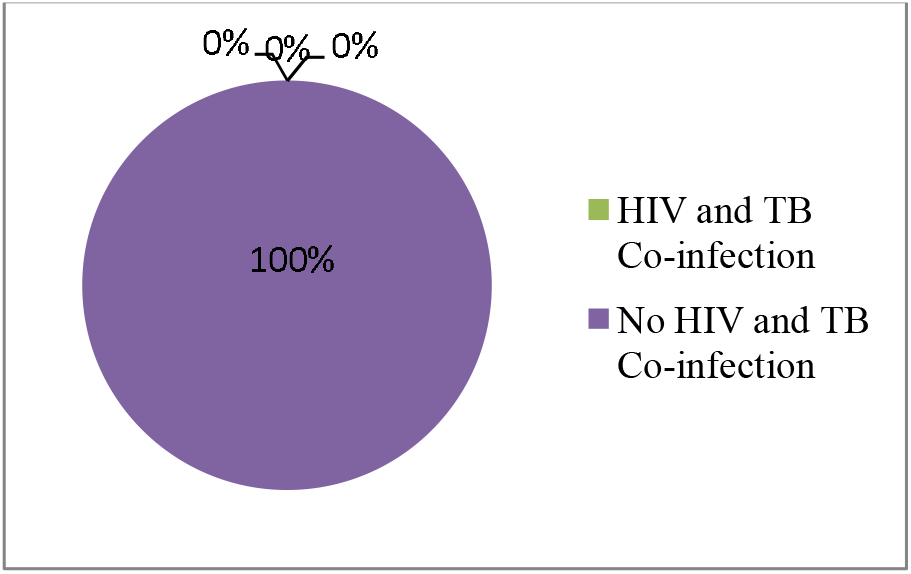
Percentage of Students with and without HIV and *Mycobacterium tuberculosis* Co-infection.

## DISCUSSION

This study was conducted to evaluate the seropositivity of HIV and *Mycobacterium tuberculosis* among University students in Port Harcourt, Nigeria. The seropositivity reported in this study showed that 2.1% were seropositive for *Mycobacterium tuberculosis* and 3.4% were seropositive for HIV while no HIV/ *Mycobacterium tuberculosis* co-infection was observed. The results of this study assumed significance and are also indicative of an emerging epidemic among University students. As the World enters the third decade of the HIV/AIDS epidemic, the evidence of its impact is undeniable, robbing countries of both human and natural resources.

This study finding revealed that the seropositivity of HIV among university students is 3.4%. This study finding is in line with the findings of White *et al*. (2009) in their study in the universities in Mali and Jembia *et al*. (2009) in their study among Cameroon undergraduates. However, the findings are slightly different from that of Helen *et al*. (2006) in their study among college students in the United States of America, with the seropositivity of 13.0%, and that of Abubakar (2008) in the Universities in Adamawa state, with seropositivity of 13.7% and that of Tobin-West *et al*. (2007) among Niger Delta students with the seropositivity of 13.7%.

The finding in the seropositivity of *Mycobacterium tuberculosis* among University students in this study shows a seropositivity rate of 2.1%. This finding corresponds with the result of Balcells *et al*. (2012), with an estimate of 2.5% and with that of Shapiro *et al*. (2012), with a rate of 2.1% students which were low seropositivity rates. Yassin *et al*. (2004) in Ethiopia, Teklu *et al*. (2012) also in Ethiopia and Nansera *et al*. (2012) in Uganda, reported in their various studies, seropositivity rates of 9.2%, 6.1% and 13.7%, respectively. These results are higher possibly as a result of the difference in the sample size, age variation of the population used for the study.

From this study, the age range of students was 16 to 39 years. *Mycobacterium tuberculosis* infection was higher in students 25 years and above. The finding that HIV seropositivity was higher in the age group 16-24 years of age in this study was consistent with other findings in Nigeria where most of the people infected with HIV were young people within the productive and reproductive age groups (Laah, 2003; National Population Commission and ICF Macro, 2009). This age group is also characterized by social vices such as teenage pregnancy, unsafe abortions, drug use and sexually transmitted infections (Laah, 2003).

In the last one-decade the seropositivity rates of the HIV and AIDS epidemic are higher in women than in men (Laah, 2003; National Population Commission and ICF Macro, 2009) as young people in Nigeria, especially women 20-24 years old, are increasingly vulnerable. In this study, the male: female ratio was kept close to 1:1. The gender-specific infection rate showed that females had a higher infection rate for HIV and *Mycobacterium tuberculosis* than their male counterparts. This high percentage recorded among the females could be attributed to riskier sexual behaviour practiced by females due to prevailing socio-cultural-economic scenarios (Okonko *et al*., 2012a). The seropositivity of *Mycobacterium tuberculosis* without HIV coinfection was also higher in females than males. The relatively higher proportion of females with HIV and AIDS (4.2%) in this study could be attributable to early age at first sex and the fact that adolescent girls tend to have older men as sex partners. This study, as well as other investigations, demonstrated that female sex was associated with most STDs, especially HIV, HBV, and HSV-2 infections (Hwang *et al*., 2000). Although high-risk sexual behaviour is much more common among young men (Bremner *et al*., 2009), data from other sources have indicated that females are disproportionately affected by HIV. In many countries like Nigeria, young women are between two to five times more likely to be infected than young men (Laah, 2003; Mamman, 2003, 2006; Panchabadeswaran *et al*., 2006; Rosen *et al*., 2008; Bremner *et al*., 2009; National Population Commission and ICF Macro, 2009; Hedden *et al*., 2009; Adebayo *et al*., 2009).

As was expected in this study, the seropositivity rate of HIV and *Mycobacterium tuberculosis* coinfection in this study was 0.0%. This means that none of the HIV seropositive students was infected with *Mycobacterium tuberculosis*. This could be owing to their level of education as HIV infection would be kept at a controlled level that no opportunistic infections such as *Mycobacterium tuberculosis* would invade the body (Jain *et al*., 2008). However, studies like that of Alex and Vivian (2012) in the central Hospital of Benin City showed a high rate of coinfection of 19.8%. Also, that of Onubogu *et al*. (2011) which was carried out in Lagos Hospitals reported 18.4% HIV and *Mycobacterium tuberculosis* coinfection and that of Obioma *et al*. (2011) in Port Harcourt City which revealed a seropositivity rate of 18.8%. Okonko et al. in 2018 had reported 14.0% among HIV-infected patients in Port Harcourt and 2020, reported 1.4% among HIV-infected patients in old Cross River State, Nigeria. This wide gap could be explained by the fact that these research works were done in hospitals and in communities where people with full-blown AIDS can be found. Also, the potential sources of variability between this study and others could be attributed in part to the analytical method or diagnostic kit used to generate the HIV and *Mycobacterium tuberculosis* data.

The limitation of this study was that sputum smear and culture was not attempted for all participants. This differs from most studies in which screening of symptoms, results of chest radiography, or both, were used as the basis for deciding which sputum specimen should be examined (Murhekar *et al*., 2004; CTCC, 2004; den Boon *et al*., 2007). The generalizability of our findings is also limited because participants were voluntary and self-selected. The power to observe statistically significant findings is limited because the study included only small numbers of cases of suspected *Mycobacterium tuberculosis* infection. The results obtained are within limits compared to similar researches (Okonko *et al*., 2012a,b,c) and also within the limits of the HIV and *Mycobacterium tuberculosis* seropositivity rate reports in Nigeria. The adult HIV seropositivity rate was reported to be 1.5% in Nigeria and 3.8% in Rivers State, Nigeria (NAIIS, 2019). This base-line data could be useful in the effective planning of tailor-made prevention and control measures among students in Port Harcourt and other similar cities. Some other limitations include that it takes a while for anti-HIV antibody to develop after infection. Anti-HIV immunoassay can therefore be false-negative. Measuring HIV RNA allows earlier detection with higher accuracy. Prior vaccination with BCG will give positive reading without active or latent BCG infection. Anti-TB immunoassay can be false-positive. Lastly, the study had limited clinical information about each patient because the information submitted to the laboratory varied and was not reliably available for inclusion in the analysis. Data about TB treatment history, patient age and sex, or HIV status are not routinely collected by all laboratories. No *M. tuberculosis* isolation was attempted.

Despite these limitations, the study provides documentation of the *Mycobacterium tuberculosis* and HIV seropositivity among students in this study area. Indeed, the convergence of *Mycobacterium tuberculosis* with the HIV epidemic may undermine gains in HIV prevention and treatment programs and requires urgent interventions. One promising way forward is the combination or integration of *Mycobacterium tuberculosis* and HIV and AIDS control programmes (Zahorka *et al*., 2004).

In conclusion, the study has revealed that the seropositivity of HIV among students in Port Harcourt (3.4%) is similar to that in other developing areas in the country, but the *Mycobacterium tuberculosis* rate was 2.1% and HIV/ *Mycobacterium tuberculosis* coinfection was zero. This does not imply a need for relaxation in the fight against these two killer diseases. Students should be advised to stop behaviour that enhances the transmission of HIV such as pre-marital sex, sharing of sharp objects such as razor blades, needles, syringes clippers, etc. New and sustained efforts must be made to contain this infection, prevent HIV transmission and expand efforts to implement ways of delivering health care to developing countries. Further studies could be undertaken to investigate other epidemiological parameters. On the side of the authority, the Government could reduce the infection rate further down by embarking on health education campaigns and training on HIV and *Mycobacterium tuberculosis* prevention.

## Data Availability

All data is contained in the body of this manuscript.

